# ARCH: Large-scale Knowledge Graph via Aggregated Narrative Codified Health Records Analysis

**DOI:** 10.1101/2023.05.14.23289955

**Authors:** Ziming Gan, Doudou Zhou, Everett Rush, Vidul A. Panickan, Yuk-Lam Ho, George Ostrouchov, Zhiwei Xu, Shuting Shen, Xin Xiong, Kimberly F. Greco, Chuan Hong, Clara-Lea Bonzel, Jun Wen, Lauren Costa, Tianrun Cai, Edmon Begoli, Zongqi Xia, J. Michael Gaziano, Katherine P. Liao, Kelly Cho, Tianxi Cai, Junwei Lu

**Author notes:** Gan and Zhou contributed equally. Cho, Cai and Lu contributed equally.

## Abstract

**Objective:** Electronic health record (EHR) systems contain a wealth of clinical data stored as both codified data and free-text narrative notes, covering hundreds of thousands of clinical concepts available for research and clinical care. The complex, massive, heterogeneous, and noisy nature of EHR data imposes significant challenges for feature representation, information extraction, and uncertainty quantification. To address these challenges, we proposed an efficient **A**ggregated na**R**rative **C**odified **H**ealth (ARCH) records analysis to generate a large-scale knowledge graph (KG) for a comprehensive set of EHR codified and narrative features.

**Methods:** The ARCH algorithm first derives embedding vectors from a co-occurrence matrix of all EHR concepts and then generates cosine similarities along with associated *p*-values to measure the strength of relatedness between clinical features with statistical certainty quantification. In the final step, ARCH performs a sparse embedding regression to remove indirect linkage between entity pairs. We validated the clinical utility of the ARCH knowledge graph, generated from 12.5 million patients in the Veterans Affairs (VA) healthcare system, through downstream tasks including detecting known relationships between entity pairs, predicting drug side effects, disease phenotyping, as well as sub-typing Alzheimer’s disease patients.

**Results:** ARCH produces high-quality clinical embeddings and KG for over 60, 000 EHR concepts, as visualized in the R-shiny powered web-API (https://celehs.hms.harvard.edu/ARCH/). The ARCH embeddings attained an average area under the ROC curve (AUC) of 0.926 and 0.861 for detecting pairs of similar EHR concepts when the concepts are mapped to codified data and to NLP data; and 0.810 (codified) and 0.843 (NLP) for detecting related pairs. Based on the *p*-values computed by ARCH, the sensitivity of detecting similar and related entity pairs are 0.906 and 0.888 under false discovery rate (FDR) control of 5%. For detecting drug side effects, the cosine similarity based on the ARCH semantic representations achieved an AUC of 0.723 while the AUC improved to 0.826 after few-shot training via minimizing the loss function on the training data set. Incorporating NLP data substantially improved the ability to detect side effects in the EHR. For example, based on unsupervised ARCH embeddings, the power of detecting drug-side effects pairs when using codified data only was 0.15, much lower than the power of 0.51 when using both codified and NLP concepts. Compared to existing large-scale representation learning methods including PubmedBERT, BioBERT and SAPBERT, ARCH attains the most robust performance and substantially higher accuracy in detecting these relationships. Incorporating ARCH selected features in weakly supervised phenotyping algorithms can improve the robustness of algorithm performance, especially for diseases that benefit from NLP features as supporting evidence. For example, the phenotyping algorithm for depression attained an AUC of 0.927 when using ARCH selected features but only 0.857 when using codified features selected via the KESER network[1]. In addition, embeddings and knowledge graphs generated from the ARCH network were able to cluster AD patients into two subgroups, where the fast progression subgroup had a much higher mortality rate.

**Conclusions:** The proposed ARCH algorithm generates large-scale high-quality semantic representations and knowledge graph for both codified and NLP EHR features, useful for a wide range of predictive modeling tasks.

## 1 Introduction

The increasing adoption of electronic health record (EHR) systems has provided opportunities for clinical studies and biomedical research ranging from patient phenotyping [2] and prediction of medical events [3], to relationship extraction between medications and adverse drug effects [4]. EHR data often cover hundreds of thousands of unique clinical features from both codified data and unstructured clinical narrative notes. With the goal of analyzing these two types of data simultaneously, the main challenges lie in combining the codified and unstructured data efficiently, representing their covered clinical features meaningfully, and quantifying statistically the presence-absence as well as the strength of relationships between different features.

The goal of combining codified and unstructured data arises from the fact that both contain clinically relevant and inextricably linked health information. Together, these complementary data sources capture a more complete picture of a patient’s medical history. The codified data, also referred to as structured data, typically consists of diagnostic codes, procedure codes, medication prescriptions, and laboratory orders and results. The utilization of codified data is straightforward; data entry is standardized and in the necessary format for analysis. For example, diagnostic codes have been used to predict the risk of heart failure [5], and procedure and medication codes have been used to predict childhood obesity [6]. Conversely, the utilization of unstructured free-text data in clinical notes is less direct [7]. This textual data covers a broad range of clinical concepts that need to be extracted via natural language processing (NLP). These NLP concepts include diseases and syndromes, clinical attributes and findings, clinical drugs, as well as laboratory, diagnostic, and therapeutic procedures, which can provide complementary information to the structured data. The NLP concepts are also referred to as clinical concept of unique identifiers (CUIs) in the Unified Medical Language System (UMLS) [8].

Many studies have shown that incorporating this textual information into analyses can enhance model performance by significant margins [9, 10]. In many cases, relevant information is only documented in clinical notes and not well codified. For instance, spontaneous reporting databases for adverse drug events are underreported when assessed using codified data only [11] since over 90% of adverse drug events are not codified [12]. As a result, it is necessary to utilize unstructured EHR data for active pharmacovigilance [13, 14]. Furthermore, NLP concepts are particularly valueable for capturing drug side effects, as a significant proportion of these effects, such as symptoms, cannot be adequately represented by diagnostic codes. For example, healthcare-associated infection (HAI), a potentially lethal condition, is widely underreported in the codified data but can be detected and even predicted using manual annotation in EHRs [15].

Combining codified and unstructured data also yields benefits for disease phenotyping. In the United States, a diagnosis code is required by the healthcare provider during the evaluation for a condition. Even if the patient is ultimately diagnosed with a different condition, the initial diagnosis code will remain in the patient’s record and may be misleading if viewed in isolation [16]. It has been shown that prediction models that combine unstructured clinical notes with codified data outperform models that utilize either unstructured or codified data alone [17, 18]. The utility of this approach is highlighted in the case of geriatric syndromes, which are associated with high morbidity, mortality, and healthcare utilization but are not fully represented by diagnostic codes found in major coding standards. Many impairments associated with geriatric syndromes, such as walking difficulty and weight loss, are not fully captured in codified fields. However, a study [19] demonstrates that incorporating unstructured data can increase the sensitivity of identifying individuals with geriatric syndromes. The supplementation of codified data with data extracted using NLP can achieve more accurate and comprehensive assessments of patient health, thereby reducing disease misclassification.

Given a large number of codified and NLP concepts, understanding their relatedness to each other can improve the efficiency of downstream predictive modeling tasks. To generate prior knowledge on the relationship among the clinical codes and NLP concepts, a potential solution is to construct a large-scale clinical knowledge graph (KG) on these concepts [20, 21, 1]. Representing EHR concepts with low-dimensional semantic embedding, KG embedding provides a quantitative glimpse into the degree of inter-relatedness of medical entities. Once high-quality embeddings of medical concepts are learned, they can improve the efficiency of downstream applications in biomedical and healthcare research including information retrieval [22, 23, 24], cohort selection [25, 26], and risk prediction [27, 28, 29].

In recent years, word embedding techniques [30, 31, 32] in NLP have been successfully applied for representing clinical concepts in a low-dimensional space. Many of these embeddings were derived for specific downstream tasks such as clustering [33] and prediction [34, 35, 3, 36]. While these embedding methods can be used to assess the relatedness of NLP concepts, they do not naturally generate a sparse KG that clearly indicates whether a link exists between entities. In addition, while KG representation techniques have been successfully used to analyze biomedical data including biomedical text and codified EHR concepts [21, 37, 38, 1, 39], joint representation of large-scale codified and NLP EHR concepts is currently lacking, as summarized in Table 1. Recently, Bai [40] proposed to jointly learn vector representations of medical concepts and words using MIMIC-III data [41]. However, their work was limited in two ways. First, they did not represent words in the clinical notes as CUIs, thus limiting the reproducibility of these representations. Second, the MIMIC-III data only contains 58, 597 in-patient visits, which confines the model performance and cannot infer broader information for outpatients. As a result, their embeddings cannot be used to generate high-quality knowledge graphs capturing general clinical information. To the best of our knowledge, there is no existing work that derives comprehensive embeddings for both codes and CUIs from a comprehensive EHR with both inpatient and outpatient data.

**Table 1:**
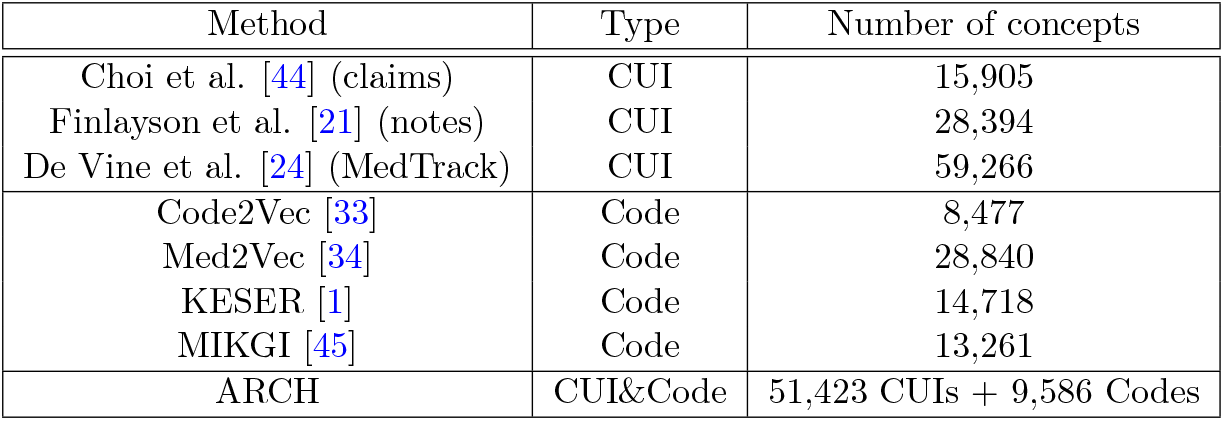
A summary of existing EHR-derived medical embeddings.

Generating KG with a large number of entities, however, is challenging for several reasons. First, an efficient computational algorithm is needed to embed all concepts when both the number of concepts and the number of EHR records are large. Second, no existing KG embedding methods provide statistical certainty on whether a link exists between two entities. Most existing KG predicts links via a supervised fashion by optimizing prediction tasks using the labeled links between entity pairs, leveraging existing knowledge of such links. While such supervised approaches can be used to assist in KG generation from EHR, it would require mapping EHR codes and narrative concepts to existing entity pairs, which itself is a challenging task. In addition, these methods necessitate the use of “negative samples”, which represent unlinked entity pairs. Unfortunately, this type of data is not readily available. Relying on the complement of positive samples as negative samples is considered unreliable, as indicated by previous research [42, 43]. These prediction-based approaches also do not provide statistical uncertainty on the existence of the link between an entity pair. Equipping the KG with certain quantification enables us to generate a sparse network while controlling for the false discovery rate (FDR).

In summary, there is a great unmet need for an approach that can integrate and summarize these high dimensional and large-scale clinical data into a KG for studies. In this paper, we will address this need by proposing an Aggregated naRrative Codified Health (ARCH) records analysis which is an efficient statistical algorithm that can generate KG embeddings along with uncertainty measures on the links. With pairwise co-occurrence counts of all EHR concepts and a few simple summary statistics, the ARCH algorithm generates low-dimensional embeddings for each concept and performs large-scale hypothesis testing based on the cosine similarity between these embedding vectors. The connectivity of entity pairs is assessed jointly by controlling for a target FDR. We validate the clinical utility of the ARCH KG, generated from EHR data from the Veterans Affairs, along with semantic embeddings through downstream tasks including detecting known relationships between entity pairs, predicting drug side effects, disease phenotyping, as well as sub-typing Alzheimer’s disease (AD) patients.

## 2 Methods

### 2.1 Generative model for the knowledge graph

Suppose there are a total of *d* EHR codified and NLP concepts, indexed by 𝒱 = {1, *…, d*}. The semantic meaning of each concept is represented by a *p*-dimensional embedding vector **V**_*j*_ for *j* = 1, *…, d*. These embeddings are generated from a latent Gaussian graphical model [46]: each column of **V** = (**V**_1_, ⋯, **V**_*d*_)^⊺^ ∈ ℝ^*d×p*^ is independent and identically distributed from *N* (0, **Θ**^−1^) where the precision **Θ** embeds the conditional dependency network of the *d* concepts, 𝒢 = (𝒱, *ε*), with the vertex set 𝒱 representing all EHR concepts and the edge set *ε* ⊆ 𝒱 × 𝒱 characterizing the conditional dependency between the concepts. Our goal is to learn the KG G with E characterized by **Θ** in that (*j, k*) ∈ ε if and only if **Θ**_*jk*_ ≠ 0 or equivalently **V**_*j*_ is conditionally dependent on **V**_*k*_ given all remaining embeddings. We aim to identify *ε* through testing the set of hypotheses ℍ = {*H*_0,*jk*_, (*j, k*) ∈ 𝒱 × 𝒱}:

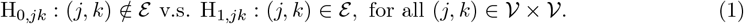

To learn the representations **V** and test ε, we assume that the observed clinical concepts in the EHR are generated from a latent Markov process driven by the embeddings sampled from the graphical model [47]. In specific, let *w*_*t*_ be the concept at time *t* and the occurrence probability of concept *j* is modeled by

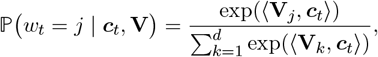

where the latent discours vector ***c***_*t*_ represents the embedding of the topic at time *t* and is generated from an autoregressive (AR) model

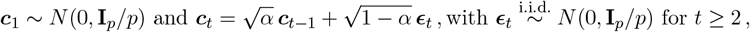

where 0 < *α* < 1 is the weight parameter. Figure 1 illustrates the generation process. The ***c***_*t*_ represents the latent topic vector at each time (e.g., phenotype, treatment, lab measurement, etc). For example, in the model part of Figure 1, ***c***_*t*_ is related to phenotype, and the probability of the concept “Alzheimer’s Disease” occurring at time *t* is larger as its embedding is closer to ***c***_*t*_. At the time *t*+1, ***c***_*t*+1_ becomes topic related to medicine and thus the concept of “Memantine” has larger occurrence probability. Under this model, the embedding inner product 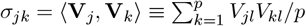 can be approximated by the population positive point-wise mutual information (PPMI) between concept *j* and *k* [48] :

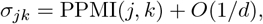

where PPMI 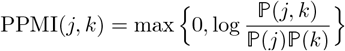, ℙ (*j, k*) is the co-occurrence probability of the concept pair (*j, k*) and ℙ (*j*) is the occurrence probability of the concept *j*. Therefore, when the number of concepts *d* has a larger order than the square root of the sample size used to estimate the PPMI, testing *H*_0,*jk*_ can be achieved by testing PPMI(*j, k*) = 0 based on the estimated PPMI.

**Figure 1.**
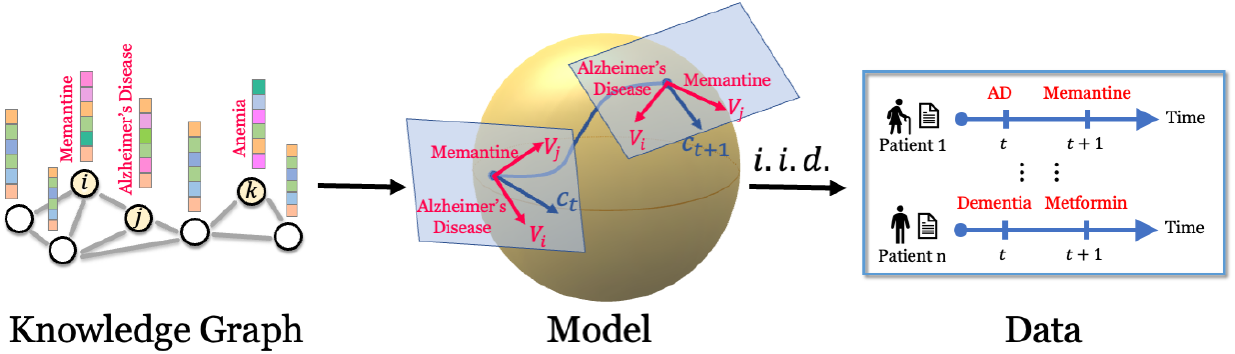
Data generation process of the EHR occurrence data. The embeddings of concepts are generated from a graphical model and the occurrence is then driven by a Markov process.

**Figure 2.**
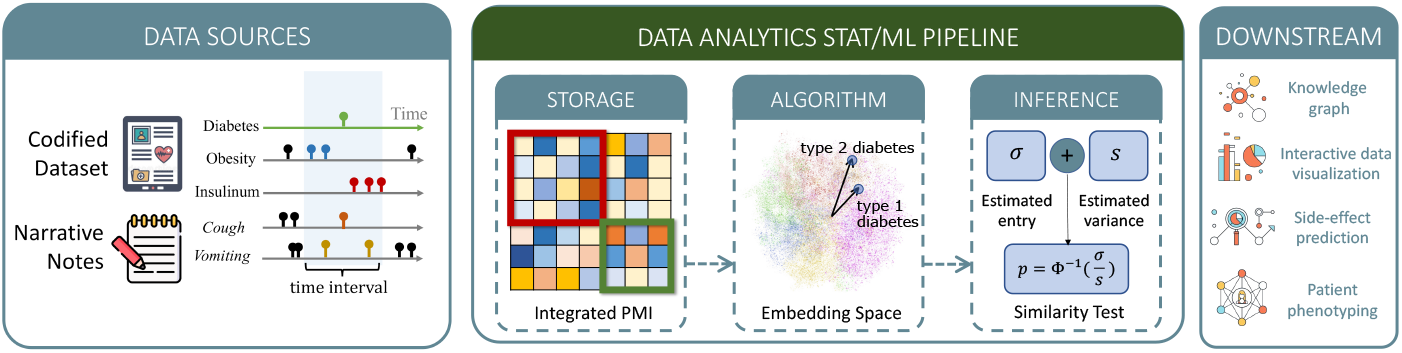
Data source, including codified data and narrative notes, and data analytics pipeline.

### 2.2 ARCH representation learning and graph recovery

For large-scale EHR datasets with a massive number of concepts and patient records, it is both statistically and computationally challenging to infer the network due to the latency and the large number of hypotheses involved. Our ARCH representation learning approach carries out the inference in two steps by (i) first screening for E by identifying marginally dependent concept pairs with nonzero pointwise mutual information, and (ii) inferring about the Gaussian graphical model structure of **Θ** via sparse regression. In the first step of screening, we apply the SURE screening [49] by selecting pairs (*j, k*) with *σ*_*jk*_ ≠ 0 after controlling for a desired FDR. In the second step, we further infer the edges from the network *G* via node-wise regression [50]. As the embedding vectors follow the Gaussian graphical model, the conditional distribution of embeddings is

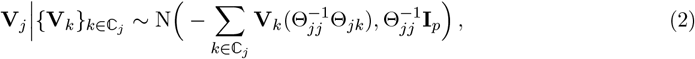

where ℂ_*j*_ is the set of concepts related to concept *j* obtained from the first prescreening step.

### 2.3 Pre-screening by PMI testing

To form a test statistic for *H*_0,*jk*_ : *σ*_*jk*_ = 0 and estimate **V**, we first calculated the empirical PPMI as ℙ ℙ 𝕄𝕝 = [ℙ ℙ 𝕄𝕝 (*j, k*)], with 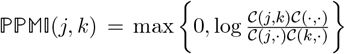, where 𝒞(*j*, ·) is the row sum of co-occurrence matrix 𝒞 (*j, k*), and 𝒞 (·, ·) is the total sum of the co-occurrence. Details for the construction of 𝒞 (·, ·) is given in Section 3.1. We next took an SVD of the empirical PPMI matrix as ℙ ℙ 𝕄𝕝 = [ℙ ℙ 𝕄𝕝 (*j, k*)] = 𝕌diag(Λ_1_, *…,* Λ_*d*_) 𝕌 ^⊺^, we can estimate **V** and population PPMI matrix of *d* concepts respectively as

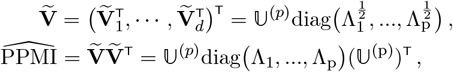

where 𝕌^(*p*)^ being the first *p* singular vectors of ℙ ℙ 𝕄𝕝 with positive eigenvalues. The dimension *p* can be selected to optimize embedding quality similar to KESER [1] by maximizing the area under the Receiver Operating Characteristics curve (AUC) of distinguishing those known relation pairs from random pairs, where known relation pairs are curated from online sources, detailed in the validation studies in Section 3.2.1.

The estimator 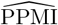 is close to the population PPMI matrix with a high approximation rate and asymptotically normal, which allows us to approximate *σ*_*jk*_ with 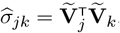. Furthermore, we may form test statistic 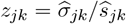 to identify *σ*_*jk*_ ≠ 0 since *z*_*jk*_ follows approximately standard normal distribution under the null hypothesis [48], where 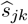 is an estimated standard error for *σ*_*jk*_ detailed in Appendix S.1. To control for multiple comparisons, we performed the Benjamini-Hochberg (BH) procedure under dependence and identified related concept pairs with *z*_*jk*_ higher than a BH-controlled threshold as detailed in Appendix S.2.

### 2.4 Sparse embedding regression

The FDR controlled testing procedure based on 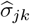 could serve as a prescreening of related concepts from the large number of concept pairs. To further screen for the most relevant concepts to form ε_*j*_ = {*k* : Θ_*jk*_ ≠ 0}, we further performed a sparse regression of 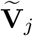 against all embedding vectors identified as related to the concept *j* after initial screening, denoted by ℂ_*j*_, to recover 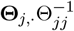 and hence its associated graph structure. Due to the potentially large number of elements identified in the pre-screening stage, we adopted an adaptive elastic-net penalized regression

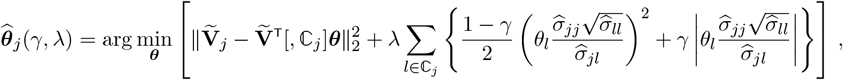

where 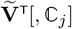 is the submatrix of 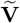 corresponding to ℂ_*j*_. The tuning parameters *λ* and *γ* control the support of 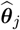 and hence the network structure. We determined the optimal values for the hyperparameters *λ* and *γ* for each target concept *j* by performing a grid search to balance the external and internal validation losses. Specifically, we computed the average of the internal Akaike information criterion (AIC) loss and an external validation loss, which was obtained using an independent dataset 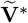, as detailed in Appendix S.3.

## 3 Validation of Real World EHR Trained ARCH Knowledge Graph

### 3.1 EHR data sources and preprocessing

We trained a large-scale ARCH KG using EHR data from the Veterans Affairs (VA) Corporate Data Warehouse (CDW), integrating both codified and narrative data from 12.6 million patients with at least 1 visit between 2000-2019. We gathered four domains of codified data including ICD diagnosis codes, procedures, lab tests, and medication prescriptions. All raw codes are rolled up to higher level codified concepts: ICD codes were aggregated into PheCodes using the ICD-to-PheCode mapping from PheWAS catalog (https://phewascatalog.org/phecodes); procedure codes, including CPT-4, HCPCS, ICD-9-PCS, ICD-10-PCS, were grouped into clinical classification software (CCS) categories based on the CCS mapping^*^; laboratory codes were either mapped to LOINC codes (https://loinc.org/) or manually annotated lab concepts; and medication codes were mapped to RXNORM codes. All free text clinical notes were processed with the Narrative Information Linear Extraction (NILE) NLP software [51], which maps clinical terms to CUIs in the UMLS. All codified and NLP data were organized as triplets: (Patient id, date, concept). Using these processed data, we created a co-occurrence matrix for all concept pairs by counting the number of co-occurrences within a 30-day window across all patients. To reduce noise, we removed concepts that have less than 3000 occurrences and concept pairs that have less than 1000 co-occurrences. Furthermore, we removed all concepts that co-occur with more than 95% of other concepts as they tend to be overly non-specific. This results in a total of over 61, 000 concepts, out of which 51, 423 are CUIs and 9, 586 are codified concepts.

### 3.2 Validation analyses

The ARCH KG was validated in four downstream tasks: (1) detecting known similar or related clinical concepts; (2) detecting drug side effects; (3) disease phenotyping; and (4) profiling of patients with AD. For the detection of known relationships and drug side effects, we also compared to embedding vectors from pretrained language model (PLM) embeddings based on Bidirectional Encoder Representations from Transformer (BERT) [52], including Self-aligning pretrained BERT (SAPBERT) [53], BERT for Biomedical Text Mining (BioBERT) [54], and BERT pretrained with PubMed (PubmedBERT) [55]. BERT’s model architecture is a multi-layer bidirectional Transformer encoder, while BioBERT, PubMedBERT and SAPBERT are pretrained on different sources based on BERT. BioBERT is pretrained on both general domain corpora and biomedical domain corpora (PubMed abstracts and PMC full-text articles), PubMedBERT is pretrained purely with in-domain text (PubMed text), and SAPBERT is pretrained on the biomedical KG of UMLS. The language model based embeddings were obtained only based on the description of the EHR concepts (e.g. preferred term for the CUI and code description).

#### 3.2.1 Detecting known relationship pairs

We curated different categories of known relation pairs from online knowledge sources including similar pairs and related pairs. Similar pairs of codified concepts were largely created based on code hierarchies including the PheCode hierarchy. Since a majority of laboratory codes in the VA are not mapped to LOINC codes, we augmented the LOINC hierarchy with manually annotated similar pairs when assessing similar laboratory code pairs. Similar CUI pairs are extracted from the relationship from the UMLS. We additionally evaluated the similarity between mapped CUI ↔ code pairs. We leveraged UMLS to obtain the mapping from different medical coding systems to concept unique identifiers [56]. For the related pairs, we first considered CUI-CUI pairs and used several categories of relationships given in the UMLS, including “<u>may treat or may prevent</u>”, “<u>classifies</u>”, “<u>differential diagnosis</u>”, “<u>method of</u>” and “<u>causative</u>”. For these CUI pairs, we map the disorder CUIs to PheCodes, drugs to the RxNorm, and procedures to CCS categories. These mapped code pairs are then further used to assess the ability to detect relatedness using codified data.

For each type of relationship, we calculated the cosine similarities of the embedding vectors of related pairs and those of randomly selected pairs to calculate AUC of the cosine similarities in distinguishing known pairs from random pairs. The random pairs were selected to match the semantic types of the related pairs. For example, when assessing “may treat or may prevent”, we restricted to disease-drug pairs. To reduce the noise of real data, we removed the features that have a pretty low frequency. Finally, we chose the dimension of embedding by optimizing the AUC. We performed ARCH testing procedure to determine whether a pair of entities are related with FDR chosen at 1%, 5%, and 10%, and reported the power of the ARCH procedure. Since no existing procedures are able to control FDR, we calculated the power of other algorithms in detecting known relationships by ranking entity pairs according to cosine similarity generated from their corresponding embeddings and then selecting the top *M* entity pairs as significant, where *M* is the number of entity pairs selected by ARCH. Among those *M* pairs, we calculated the proportion of those known to be related as their power.

#### 3.2.2 Detecting drug side effect

The unintended effects or adverse events (AEs) of drugs threaten public health and patient safety [57]. However, the screening for and adjudication of AEs is costly and time-intensive and post-market drug retraction is expensive [58]. It is thus critical to predict the potential AEs of drugs prior to their widespread use. The ARCH KG provides semantic representations for both drugs and side effects, which can be subsequently modeled to identify potential side effects for a given drug. ARCH network includes both narrative and codified features, which can improve our ability to detect side effects that tend to be under-codified in the EHR. To develop and validate a side effect prediction model based on ARCH embeddings, we obtained labels from the Side Effect Resource (SIDER)^†^ database of drugs and adverse drug reactions (ADRs) [59]. The SIDER database captures side-effect information from multiple data sources including placebo-controlled clinical trials, the FDA Adverse Event Reporting System (AERS), and biomedical literature. We followed the data cleaning procedure outlined in multimodal representation learning [60] and selected common AEs reported in more than 50 drugs. The AEs were mapped to both PheCodes and CUIs while the drugs, recorded as DrugBankID in SIDER, were mapped to RxNorm codes and CUIs. Following these steps using the VA data, we obtained 831 drugs and corresponding 4, 010 AEs, which compose in total 128, 220 drug-AE pairs. Similar to relation detection, we randomly sampled the same number of negative pairs from those drug-disease entity pairs that have not been reported as drug-AE pairs. The AUC and power for detecting drug side effects based on ARCH embeddings or *p*-values as well as based on embeddings from existing language models were calculated similarly as those for the relation detection. Since the drug-AE pairs can exist in four forms: RxNorm-CUI pairs, CUI-CUI pairs, RxNorm-PheCode pairs, and CUI-PheCode pairs, we took the highest score among these four relationship pairs to represent the final score for each drug-AE pair. We also compared the score that uses all four forms of data to the score based on codified data only, i.e. RxNorm-PheCode pairs, with respect to their power in detecting the drug-AE pairs. Since this KG representation can be viewed as a pre-training step that can be further fine-tuned for the task of AE detection, we further evaluated the quality of ARCH embeddings as well as embeddings from existing language models based on the performance of a few-shot supervised model for this task. The fine-tuning step employed a commonly used loss function [61] as detailed in Appendix S.5. We used 1% of the positive and negative pairs to estimate model parameters, another 1% as validation data to select optimal tuning parameters, and the remaining 98% pairs as a test data set for evaluation.

#### 3.2.3 Disease phenotyping

A major bottleneck for conducting translational research studies with EHR is the lack of large-scale precise data on disease outcomes needed for predictive modeling. For most conditions, ICD codes do not accurately reflect the true disease status while manual annotation via chart review is not scalable [62]. Recently, many unsupervised machine learning based phenotyping algorithms have been shown to greatly improve the case definition over ICD codes [63, 64, 65, 62, 66]. However, most of these algorithms require the specification of relevant features. Given the large number of potential EHR features, automatically selecting features important for a disease of interest is an important step to ensure the accuracy of the downstream modeling. We next illustrate how the ARCH network can serve as an effective feature selection tool for EHR phenotyping and compare to the existing KG based feature selection tool, KESER [1], which only identifies codified features. To compare the performance of ARCH versus KESER, we employed the unsupervised PheNorm algorithm [65]. PheNorm can be viewed as weakly supervised in that it treats the counts of the PheCode and/or CUI corresponding to the disease as “silver standard labels” to train an algorithm that combines these key features with additional informative features including a measure of healthcare utilization via drop-out training and mixture modeling. We compared PheNorm trained with ARCH selected features, PheNorm trained with only KESER selected features, the MAP algorithm which only uses counts of the main PheCode and CUI, and healthcare utilization [62], as well as two benchmark methods that use the logarithm of the count of the main disease ICD code plus one (Main ICD Only) and the logarithm of the count of the mention of the disease CUI plus one (Main NLP Only) as the disease predictive scores, respectively. Since KESER only includes codified features and MAP only uses the three key features, these comparisons also illustrate the value of other informative features, particularly NLP features from free text, in improving the accuracy of the algorithm. We trained theses phenotyping algorithms using EHR data from 53, 549 MGB Biobank participants for 8 conditions: coronary artery disease (CAD), Crohn’s disease (CD), rheumatoid arthritis (RA), ulcerative colitis (UC), Congestive heart failure (CHF), type 1 diabetes mellitus (T1DM), type 2 diabetes mellitus (T2DM) and depression. To evaluate their accuracy, the CAD, CD, RA, UC, CHF, T2DM, T2DM and depression phenotyping algorithms were validated against 187, 138, 154, 127, 114, 540, 285 and 540 labeled observations curated via manual chart review, and the AUCs were reported.

#### 3.2.4 Profiling of AD patient via ARCH embeddings

Semantic representation of the EHR concepts can be linked with patient level EHR data to represent patient clinical profile [67, 68, 69]. These patient embeddings can then be applied to perform downstream tasks such as identifying “*patient like me”* [70] and mortality prediction [71]. However, representing a patient’s clinical profile with respect to a specific condition, such as AD, requires the knowledge of other EHR features relevant to AD progression as well as their relative importance [72]. Our ARCH KG serves this purpose in that it can generate embeddings to represent an AD patient. To demonstrate this, we used EHR data of 38,267 patients with AD diagnosis, collected from the University of Pittsburgh Medical Center (UPMC) over the period 2011-2021. We selected the AD relevant features and generate embeddings for the *i*th patient using the following term frequency–inverse document frequency (TF-IDF) procedure:

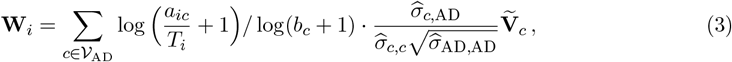

where 𝒱_AD_ is the feature set related to AD detected by ARCH, *T*_*i*_ is the follow-up time of the *i*th patient, 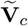 is the estimator of word representation for concept *c, a*_*ic*_ is the occurrence of the feature *c* in the EHR of the *i*th patient, *b*_*c*_ is the occurrence of feature *c* in all patients from VA between 2000-2019. Together, the PMI testing procedure and clinical embeddings can help us to generate patient embeddings that present phenotyping. As an illustration, we applied *k*-means algorithm to cluster patients into two groups using the patient embeddings. We analyzed the mortality risk of the two groups using the Kaplan-Meier (KM) curve of the time from first AD diagnosis to death. We characterized the between group differences in patient profile with respect to the distributions of AD related features selected via ARCH. For each AD related feature within each group, we compute its average intensity defined as the concept count normalized by total feature count within each patient. We summarize the group difference in patient profile based on the between-group differences in feature intensity.

## 4 Results

By optimizing the AUC of distinguishing known relation pairs from random pairs as detailed in Section 3.2.1, we set the dimension of embeddings as *r* = 1500 to optimize the embedding quality. We worked with 1500-dimensional embeddings on the following tasks.

### 4.1 Detecting known relationship pairs

The AUCs and power in detecting known relationships are summarized in Table 2 with details on the accuracy of detecting specific types of relationships given in Table 5 in Appendix S.6. The embeddings trained by ARCH achieved an AUC of 0.871 for detecting similar pairs and 0.836 for detecting related pairs, while pretrained language model derived embeddings including Pubmed-BERT, BioBERT and SAPBERT attained much lower AUCs ranging from 0.583 to 0.735. The ARCH screening procedure attained power of 0.909 for similar pairs 0.892 for related pairs under the target FDR 0.1, while the highest power among the three benchmarks was only 0.74 for similar pairs and 0.70 for related pairs. Visualizations of the ARCH network can be found at https://celehs.hms.harvard.edu/ARCH/, which enables users to visualize concepts relevant to a set of target concepts.

**Table 2:** AUCs and power of detecting known similar pairs and related pairs with different algorithms with various target FDRs. Pub stands for PubmedBERT, Bio stands for BioBERT and SAP stands for SAPBERT.

|  | FDR | type | ARCH(p) | ARCH(c) | Pub | Bio | SAP |
| --- | --- | --- | --- | --- | --- | --- | --- |
| AUC |  | similar | **0.873** | 0.871 | 0.670 | 0.589 | 0.735 |
|  |  | related | 0.832 | **0.836** | 0.649 | 0.583 | 0.642 |
| Power | 0.1 | similar | **0.909** | 0.902 | 0.677 | 0.548 | 0.741 |
|  |  | related | **0.892** | 0.884 | 0.701 | 0.594 | 0.672 |
|  | 0.05 | similar | **0.906** | 0.898 | 0.668 | 0.536 | 0.733 |
|  |  | related | **0.888** | 0.880 | 0.691 | 0.583 | 0.659 |
|  | 0.01 | similar | **0.900** | 0.892 | 0.646 | 0.514 | 0.715 |
|  |  | related | **0.880** | 0.871 | 0.670 | 0.559 | 0.638 |

### 4.2 Identifying drug side effects

Table 3 shows the AUC-ROC and power of ARCH embeddings, the pre-trained language model embeddings, as well as the *p*-values from ARCH screening testing procedures in detecting drug side effects. The unsupervised ARCH embeddings and the screening test *p*-values achieved substantially a higher AUC of 0.723 and 0.747, compared to those from PLM which ranged from 0.584 to 0.634. With few-shot supervised training, the ARCH embeddings attained an AUC of 0.826 while the AUC of the fine-tuned PLM embeddings remained below 0.69. Comparing the power in detecting drug side effects using codified data alone versus both codified and NLP data, we find that adding NLP information greatly improved the ability to capture side effects for most drug classes as shown in Figure 3. In Figure 4, we show most of the side effects of Levothyroxine and Hydrocodone can be detected by ARCH while a significant fraction of the side effects can only be captured with the help of NLP data. More examples of word-cloud figures are shown in Figure 10 in Appendix S.6.

**Table 3:**
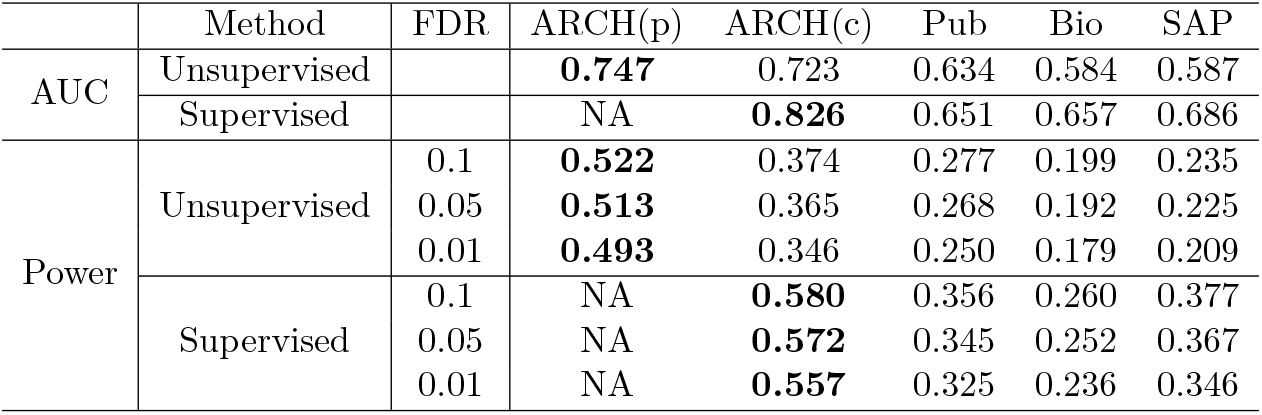
AUCs and the sensitivities of different benchmark methods compared with ARCH for identifying drug side effects. The first and the third blocks show the performance of each method without supervision, while the second and the fourth blocks show the performance of the method with supervised learning using 1% drug-side effects pairs for training. Pub stands for Pubmed-BERT, Bio stands for BioBERT and SAP stands for SAPBERT.

**Figure 3.**
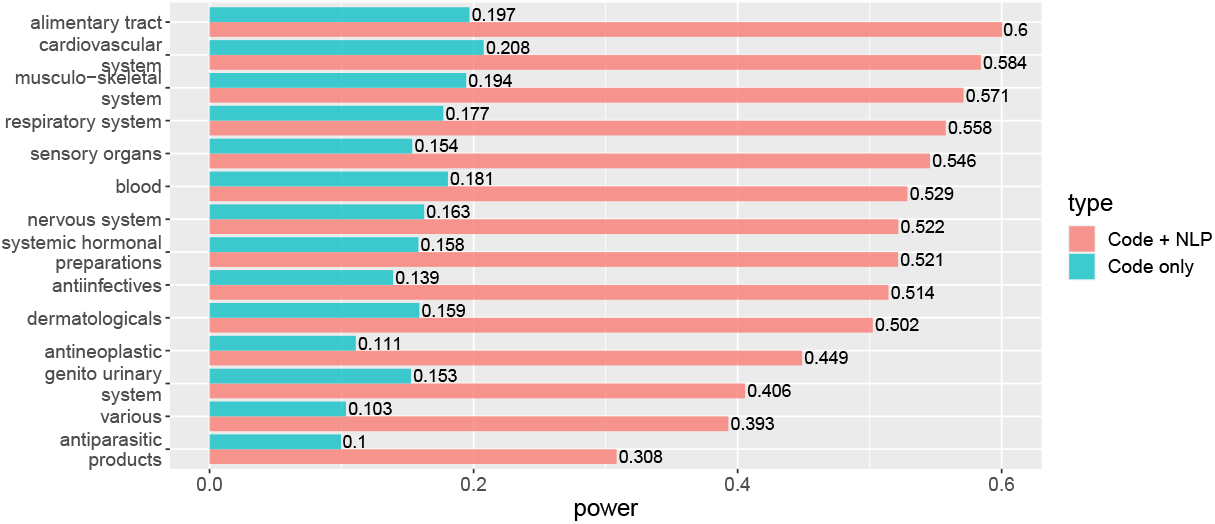
Sensitivity of detecting drug-side effects pairs with only codified data and that with both codified data and NLP with ARCH under target FDR 0.05.

**Figure 4.**
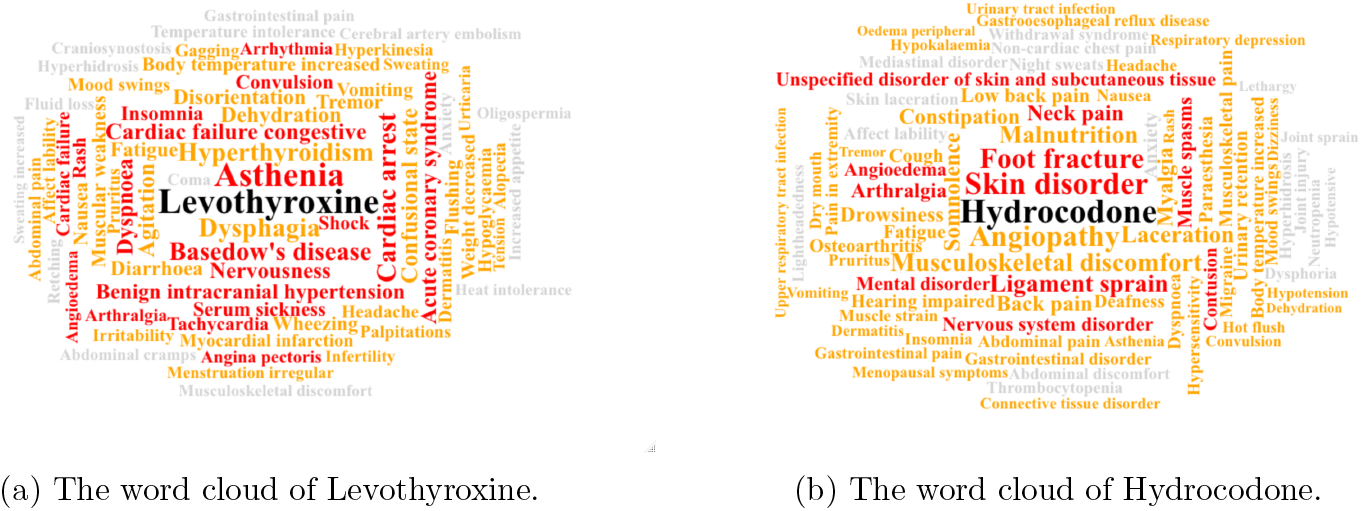
The word clouds of the side effects of two sample drugs - (a) Levothyroxine on the left and (b) Hydrocodone on the right. The surrounding words describe side effects. The words colored red are detected using codified data only while the words colored orange or red are detected by using both codified data and NLP codes. The words colored by grey are undetected. The size of the words is determined by the cosine similarity with the target drug code.

### 4.3 Disease phenotyping

Figure 5 shows the AUCs of 8 phenotyping algorithms validated on labeled data from MGB. PheNorm with ARCH selected features performs the best among all methods. The AUCs of the PheNorm algorithms with features selected by ARCH exceeded 0.9 for all 8 diseases and on average were 0.028 (*p*-value 3.30 × 10^*−5*^), 0.067 (*p*-value 9.87 × 10^*−12*^), 0.081 (*p*-value 3.29 × 10^*−11*^), and 0.076 (*p*-value 1.06 × 10^*−11*^) higher than that of PheNorm with KESER features, MAP, ICD only and NLP only. The gain in performance is particularly noteworthy for conditions that benefit from NLP features. For example, after applying ARCH in the feature selection step, the AUC of the PheNorm algorithm for depression increased from 0.857 of KESER to 0.927 (*p*-value 2.47 × 10^*−4*^).

**Figure 5.**
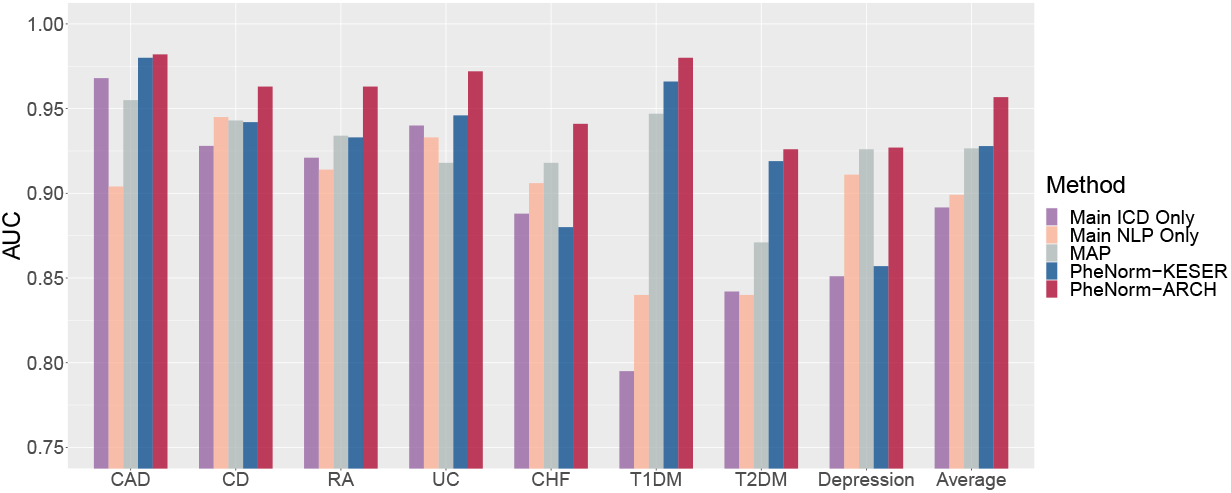
The AUC of different phenotyping algorithms trained with different feature sets across 8 diseases.

### 4.4 Profiling of AD patient via ARCH embeddings

The AD cohort consists of about 64.7% female patients, 90.3% white and 7.6% black patients, with an average age of 82 years at first ICD code for AD and an average lifespan of 86 years. K-means clustering of the ARCH-based patient embeddings as detailed in Section 3.2.4 resulted in two subgroups: a fast progression group consisting of 12.3% the patients and a slow progression group formed by the remaining patients. As shown in Figure 6, the 5-year survival rate was 42.0% (95% CI: [38.6%, 45.7%]) and 80.9% (95% CI: [80.3%, 81.6%]) for the fast and slow progression groups, respectively.

**Figure 6.**
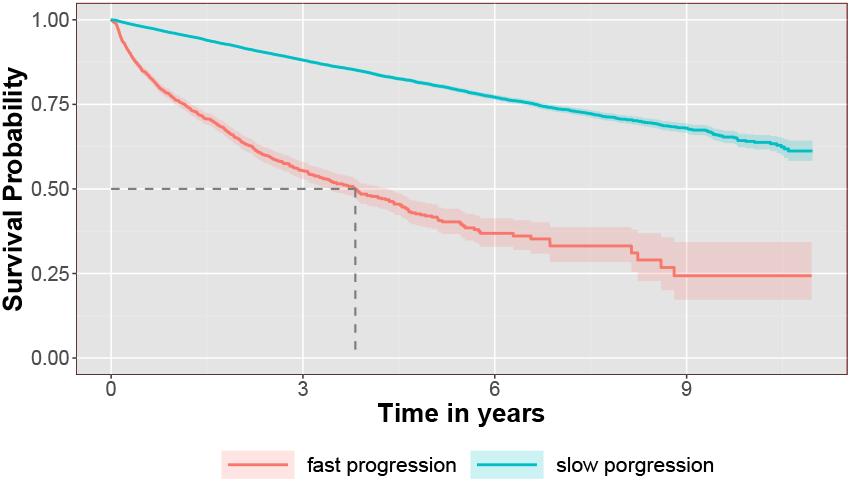
The KM survival curves for the fast and slow progression groups identified via *k*-means clustering of the ARCH patient level embeddings.

Figure 7 highlights the top disease and drug features with the largest differences between the fast and slow progression groups. The phenotype features associated with faster progression are common phenotypes at the late stage of AD. Pneumonia is one of the two most serious medical conditions seen in late-stage AD patients [73]; hypovolemia and hypernatremia may be found in association with dehydration, which can occur in impaired late-stage AD patients who are dependent on others for fluid intake [74, 75, 76]. On the other hand, the features that appear more frequently in the slow progression group of patients, which are colored blue in the figure, are either common signs or possible causes of AD. Memory deficits begin from the early stage of AD [77], while vitamin deficiency and hypothyroidism are risk factors for AD [78, 79, 80]. As shown in the network of drug features and procedure features, the features ‘atorvastatin’, ‘metformin’, ‘escitalopram’, ‘melatonin’, among others, have been shown to moderate AD or slow down the progression of cognitive impairment in AD patients [81, 82, 83, 84]. Memantine, a type of N-methyl-D-aspartate receptor antagonist, is the only drug approved for use in moderate to severe AD under current AD treatment guideline [85, 86]; Rivastigmine and Donepezil are the drugs approved by FDA (Food and Drug Administration) for AD treatment besides Memantine and two accelerated approval drugs^‡^; all these three drugs are more common in the fast progression group of patients. With these references, the clustering of patients is practical and realistic, indicating the good quality of patient embedding based on the feature selection by ARCH.

**Figure 7.**
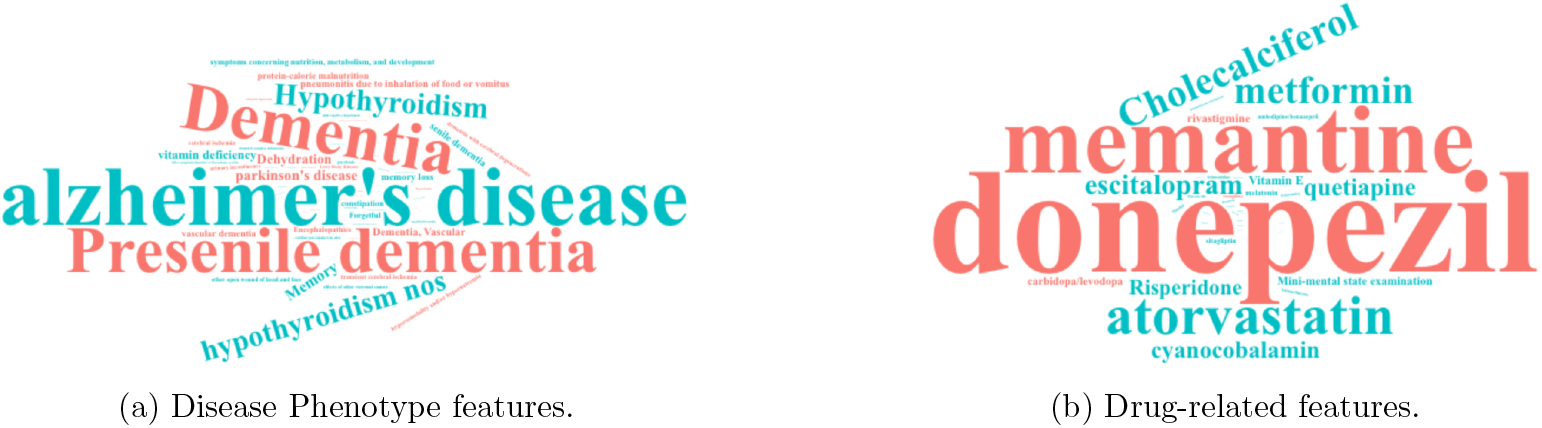
The word cloud of (a) phenotype features; and (b) drug features that drive the differences between the two subgroups. The size of the feature is determined by the between-group difference in the average intensity of such a feature. Red-colored features represent higher average intensity in the fast progression group and blue-colored features represent higher intensity in the slow progression group.

**Figure 8.**
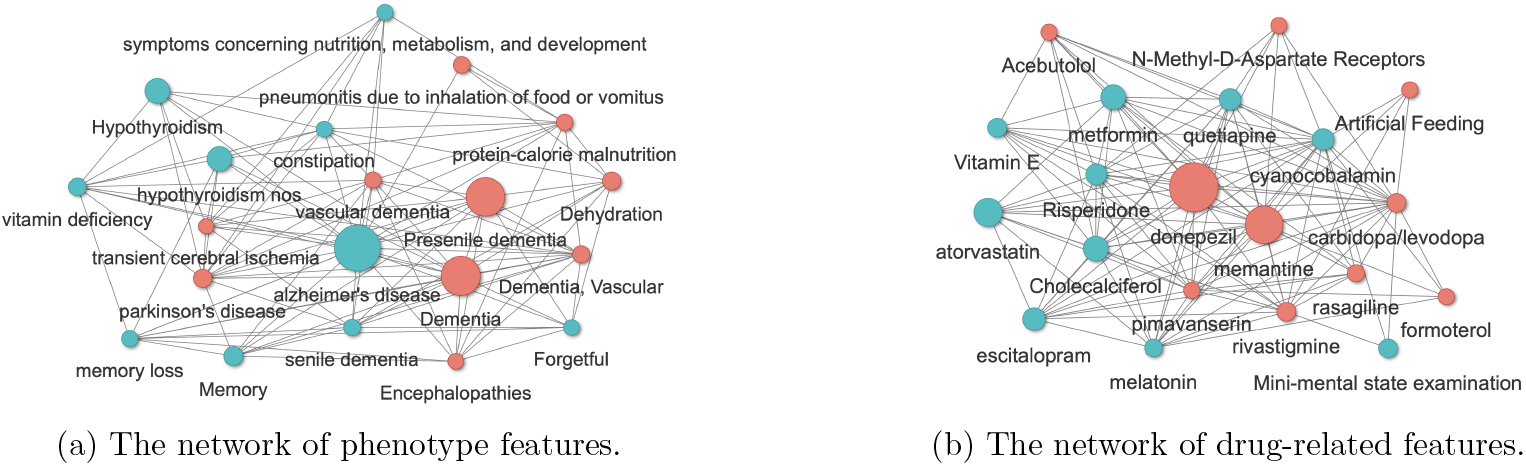
The network of (a) phenotype features; and (b) drug features that drive the differences between the two subgroups. The size of the feature is determined by the between-group difference in the average intensity of such a feature. Red-colored features represent higher average intensity in the fast progression group and blue-colored features represent higher intensity in the slow progression group.

## 5 Discussion

Utilizing summary-level EHR data, the ARCH KG learning approach provides a highly scalable method for effectively representing codified and narrative EHR concepts on a large scale, while also recovering their network structure. The VA EHR-derived ARCH embeddings represent the first large-scale EHR embeddings to include both codified and NLP concepts, with the incorporation of NLP concepts proving particularly beneficial in real-world applications such as drug side effect monitoring and disease phenotyping. Additionally, the network structure derived from ARCH is constructed with a statistically guaranteed false discovery rate.

The versatility of the learned ARCH embeddings makes them ideal for a broad range of down-stream tasks. These embeddings demonstrate greater robustness than existing PLM-based embeddings. Our semantic representation evaluations and drug side effect prediction studies show that the ARCH embeddings can effectively capture the semantic relationships between EHR entities and concepts. Our results indicate that the ARCH embeddings with few shot training have the potential to achieve high accuracy in KG-related tasks, such as entity matching and relation extraction. Additionally, the ARCH embeddings can serve as pre-trained representations of EHR concepts that can be linked to individual-level EHR data, further improving patient-level prediction tasks, as demonstrated in the AD patient profiling study. Joint representations of both codified and NLP data also enable more comprehensive multi-modal modeling of EHR data, significantly enhancing prediction performance for outcomes that require predictors that are not well-coded.

The use of summary-level data in learning the ARCH network creates an opportunity for collaborative training of knowledge graphs across multiple institutions. This approach can enhance the quality of the trained representation and improve the portability of downstream prediction algorithms. However, co-training ARCH embeddings using multi-institutional data faces a challenge in dealing with coding differences between institutions. Even for institutions that have mapped their local EHR codes to a common ontology, such mappings are often incomplete. Future research needs to explore co-training knowledge graphs for overlapping yet non-identical EHR concepts from multiple institutions based on summary-level data.

Currently, the ARCH network relies solely on EHR occurrence patterns of concepts, disregarding valuable information contained in their descriptions. Incorporating both occurrence patterns and descriptions through language models is an intriguing avenue for further research in improving the network.

## Data Availability

The data that support the findings of this study are available from the Veterans Affairs (VA) but restrictions apply to the availability of these data, which were used under license for the current study, and so are not publicly available. Data are however available from the authors upon reasonable request and with permission of VA.

https://celehs.hms.harvard.edu/ARCH/

## Acknowledgements

We would like to acknowledge the invaluable contributions arising from the collaboration between Veterans Affairs (VA) and the Department of Energy (DOE) which provided the computing infrastructure essential to develop and test these approaches at scale with nationwide VA EHR data. This project was supported by the NIH grants 1OT2OD032581, R01 HL089778 and R01 LM013614, P30 AR072577, and the Million Veteran Program, Department of Veterans Affairs, Office of Research and Development, Veterans Health Administration, and was supported by the award #MVP000. This research used resources from the Knowledge Discovery Infrastructure at Oak Ridge National Laboratory, which is supported by the Office of Science of the US Department of Energy under Contract No. DE-AC05-00OR22725. This publication does not represent the views of the Department of Veterans Affairs or the U.S. government.

## Competing Interests

The authors declare that there are no competing interests.

## Author Contribution

ZG: Methodology, Software, Writing – original draft. DZ: Methodology, Software, Writing – original draft. ER: Resources. VAP: Data curation. YH: Data curation. GO: Resources. ZX: Methodology. SS: Writing - review & editing. XX: Visualization. KFG: Writing - review & editing. CH: Writing - review & editing. CB: Visualization. JW: Data curation. LC: Writing - review & editing. TC: Writing - review & editing. EB: Writing - review & editing. ZX: Writing - review & editing. JMG: Writing - review & editing. KPL: Writing - review & editing. KC: Conceptualization, Writing – review & editing, Supervision. TC: Methodology, Conceptualization, Writing – review & editing, Supervision. JL: Methodology, Conceptualization, Writing – review & editing, Supervision, Project administration, Funding acquisition.

## Footnotes

* https://www.hcup-us.ahrq.gov/toolssoftware/ccs_svcsproc/ccssvcproc.jsp

† http://sideeffects.embl.de

‡ https://stanfordhealthcare.org/medical-conditions/brain-and-nerves/alzheimers-disease/treatments/medications.html

## Notes

### Competing Interest Statement

The authors have declared no competing interest.

### Author Declarations

Ethics committee/IRB of the Veterans Affairs gave ethical approval for this work

