## Supplemental for "ARCH: Large-scale Knowledge Graph via Aggregated Narrative Codified Health Records Analysis"

*Technical supplementary material to*

This document contains the supplementary material to the paper "ARCH: Large-scale Knowledge Graph via Aggregated Narrative Codified Health Records Analysis".

### S.1 Estimation of variance of PMI matrix

To estimate the standard deviation of $\widetilde{\mathbf{V}}_i^\intercal \widetilde{\mathbf{V}}_j$, denoting this estimator as $\widehat{\sigma}_{ij}$, we assume the following model:

$$X_{i,w}(t) \overset{i.i.d}{\sim} \text{Bernoulli}(1, p_w) \ \text{ for } 1 \le w \le d, 1 \le t \le T, 1 \le i \le n\,,$$

where $X_{i,w}(t)$ is the indicator of the occurrence of concept $w$ at time $t$ for the $i$-th individual, and $p_w$ is the marginal probability for the occurrence of concept $w$. This variance estimator is advantageous as it avoids the need to use patient-level data or apply the bootstrap algorithm, which can be computationally expensive and time-consuming. Furthermore, sharing patient-level data may not be feasible due to privacy and security concerns. Thus, our model is scalable, computationally economical, and avoids administrative challenges, making it easier to comply with privacy regulations.

Denote $\mathbf{E} = \mathbb{PPMI} - \mathbb{PPMI}^*$, $\mathbf{P} = \mathbb{U}^*_{n\times n}(\mathbb{U}^*_{n\times n})^\intercal$, $\widehat{\mathbf{P}} = \mathbb{U}^{(r)}(\mathbb{U}^{(r)})^\intercal$, $\mathbf{M}$ as the difference of low-rank estimator of PPMI and true PPMI, where $\mathbb{PPMI}^*$ is true PPMI matrix and $\mathbb{U}^*$ is the sigular vectors of true PPMI matrix. Denote $T$ as the average number of concepts per patient's health record, $q$ as the average number of concepts in one window size of one patient's health record, $p_i = \dfrac{\mathcal{C}(i,\cdot)}{\sum_{k=1}^n \mathcal{C}(k,\cdot)}$ as the marginal proportion for feature $i$. Then the estimated variance of the $(i,j)$th entry of the low-rank PPMI matrix can be computed with below equation:

$$\begin{aligned}
\widehat{\text{Cov}}(\mathbf{M}_{ij}) \approx & \widehat{\text{Cov}}\Big(\mathbf{e}_j^\intercal(\mathbf{PE} + \mathbf{EP})\mathbf{e}_i\Big) \\
= & \Big(\mathbf{P}\widehat{\text{Cov}}(\mathbf{E}_{\cdot i})\mathbf{P}\Big)_{j,j} + \sum_{k=1}^n \mathbf{P}_{ki}^2\Big(\widehat{\text{Cov}}(\mathbf{E}_{\cdot k})\Big)_{j,j} + \sum_{1\le k\ne l\le n} \mathbf{P}_{ki}\mathbf{P}_{li}\Big(\widehat{\text{Cov}}(\mathbf{E}_{\cdot k}, \mathbf{E}_{\cdot l})\Big)_{j,j} + \\
& \sum_{k=1}^n \Big\{\mathbf{P}_{ki}\Big(\widehat{\text{Cov}}(\mathbf{E}_{\cdot k}, \mathbf{E}_{\cdot i})\mathbf{P}\Big)_{j,j} + \mathbf{P}_{ki}\Big(\mathbf{P}\widehat{\text{Cov}}(\mathbf{E}_{\cdot i}, \mathbf{E}_{\cdot k})\Big)_{j,j}\Big\}.
\end{aligned} \tag{S.1}$$

Denote $T_1 = Tq - \dfrac{q(q+1)}{2}$, we have:

$$\begin{aligned}
\widehat{\text{Cov}}\big(\mathbf{E}_{i\cdot}\big) &= \frac{1}{nT_1p_i}\big(\mathbf{1}\mathbf{1}^\top(p_i - \frac{1}{2}) - \frac{1}{2}\mathbf{1}\mathbf{e}_i^\top - \mathbf{e}_i\mathbf{1}^\top\frac{1}{2} + \text{diag}(p_j^{-1})\frac{1-p_i}{2} + \frac{1}{2\boldsymbol{p}_i}\mathbf{e}_i\mathbf{e}_i^\top\big)\,, \\
\widehat{\text{Cov}}\big(\mathbf{E}_{i\cdot}, \mathbf{E}_{j\cdot}\big) &= \frac{1}{nT_1}\big(\mathbf{1}\mathbf{1}^\top - \frac{1}{2p_i}\mathbf{1}\mathbf{e}_i^\top - \frac{1}{2p_j}\mathbf{e}_j\mathbf{1}^\top - \frac{1}{2}\text{diag}(p_j^{-1}) + \frac{1}{2p_ip_j}\mathbf{e}_j\mathbf{e}_i^\top\big)\,.
\end{aligned} \tag{S.2}$$

Once the entry of PPMI and the variance of the entry of PPMI are estimated, denoted by $\mathbb{PPMI}(i,j)$ and $\widehat{\text{Cov}}(\mathbb{PPMI}(i,j))$, the $p$-value for testing whether $\mathbb{PPMI}^*(i,j)$ equals to zero from $z$-test is $p_{ij} = 1 - \Phi\big(\mathbb{PPMI}(i,j)/\widehat{\text{Cov}}(\mathbb{PPMI}(i,j))\big)$, where $\Phi$ is the cumulative distribution function (CDF) of a standard normal distribution.

### S.2 BH procedure under dependence

We have $n$ hypotheses, $H_{0,i}$ and $p$-values $p_i$ for each. Assume that under $H_{0,i} : p_i \sim U(0;1)$. First we order $n$ $p$-values $p_{(1)} \leq p_{(2)} \leq \cdots \leq p_{(n)}$ and let $H_{(1)}, H_{(2)}, \cdots, H_{(n)}$ be the corresponding hypotheses. Then we compute $k = \max_i\{i : p_{(i)} \leq \frac{i}{n(\ln(n)+1}\alpha\}$, where $\alpha$ is the target FDR. Finally, we reject all $H_{(i)}, i \leq k$.

### S.3 Parameter tuning for elastic net regularization

To obtain test residual, we obtained cooccurrence matrix from a different constitution and construct its word representation $\widetilde{\mathbf{V}}^*$ in the same way above, and aligned $\widetilde{\mathbf{V}}$ and $\widetilde{\mathbf{V}}^*$ so that the $i$th row of $\widetilde{\mathbf{V}}$ represents the same concept as the $i$th row of $\widetilde{\mathbf{V}}^*$. If the concept did not exist in the second institution, we filled the corresponding row of $\widetilde{\mathbf{V}}^*$ with zeros. Then we computed

$$
\begin{aligned}
\lambda_j^{\mathrm{A}} &= \arg\min_{\lambda} \left( \ln\left(\|\widetilde{\mathbf{V}}_j - \widetilde{\mathbf{V}}[,\mathbb{C}_j]\widehat{\boldsymbol{\theta}}_j(\gamma,\lambda)\|\right) + \frac{N_j(\gamma,\lambda)}{p} \right), \\
\lambda_j^{\mathrm{B}} &= \arg\min_{\lambda} \left( \ln\left(\|\widetilde{\mathbf{V}}_j^* - \widetilde{\mathbf{V}}^*[,\mathbb{C}_j]\widehat{\boldsymbol{\theta}}_j(\gamma,\lambda)\|\right) + \ln\left(\|\widetilde{\mathbf{V}}_j - \widetilde{\mathbf{V}}[,\mathbb{C}_j]\widehat{\boldsymbol{\theta}}_j(\gamma,\lambda)\|\right) + \frac{N_j(\gamma,\lambda)}{p} \right), \\
\lambda_j &= \begin{cases} \min\{\lambda_j^{\mathrm{A}}, \lambda_j^{\mathrm{B}}\} & , \text{ if } \mathbf{V}_j^{*\intercal}\mathbf{V}_j^* \neq 0 \\ \lambda_j^{\mathrm{A}} & , \text{ if } \mathbf{V}_j^{*\intercal}\mathbf{V}_j^* = 0 \end{cases}
\end{aligned}
$$

where $\{\lambda_j \mid j = 1, \cdots, d\}$ are the final parameters we selected for elastic net regularization.

### S.4 Data source

| | | | CUI | | | | |
|---|---|---|---|---|---|---|---|
| Semantic type | ACTI | CHEM | DISO | PHEN | PHYS | PROC | Total |
| Number | 353 | 12828 | 28282 | 1081 | 515 | 8364 | 51423 |
| | | | Codes | | | | |
| Class | CCS | Lab | PheCode | RxNorm | | | Total |
| Number | 224 | 6025 | 1776 | 1561 | | | 9586 |

Table 4: Number of features in each category.

### S.5 Supervised learning for identifying drug side effects

The loss function for the supervised learning is defined below:

$$
\begin{aligned}
\mathcal{L}(M) &= \frac{1}{\alpha}\sum_i \log\left(1 + \sum_{j\in\mathcal{P}_i} \exp\left(-\alpha(S_{ij}-\lambda)\right)\right) + \frac{1}{\beta}\sum_i \log\left(1 + \sum_{j\in\mathcal{N}_i} \exp\left(\beta(S_{ij}-\lambda)\right)\right) \quad , \\
\frac{\partial\mathcal{L}}{\partial M} &= -\sum_i \sum_{j\in\mathcal{P}_i} \frac{\mathbf{x}_i\mathbf{x}_j^\intercal \exp(-\alpha S_{ij})}{\exp(-\lambda\alpha) + \sum_{j\in\mathcal{P}_i}\exp\left(-\alpha S_{ij}\right)} + \sum_i \sum_{j\in\mathcal{N}_i} \frac{\mathbf{x}_i\mathbf{x}_j^\intercal \exp(\beta S_{ij})}{\exp(\lambda\beta) + \sum_{j\in\mathcal{N}_i}\exp\left(\beta S_{ij}\right)} \quad ,
\end{aligned}
$$

where $S_{ij} = \mathbf{x}_i^\intercal M \mathbf{x}_j$, $\mathbf{x}_i$ is the embedding vector of feature $i$, computed by $\mathbf{x}_i = \mathbf{V}_i \widehat{\sigma}_{ii}^{-1}$, $\mathcal{P}_i$ is the set of side effects related with the $i$th drug in the training data set, and $\mathcal{N}_i$ is the set of phenotypes unrelated with the $i$th drug in the training data set. Once $M$ is obtained by minimizing the loss function above, we define $S_{ij} = \mathbf{x}_i^\intercal M \mathbf{x}_j$ as the score of the pairs of drug $i$ and side effects $j$. We then computed the AUC on the validation dataset to select the optimal parameters for each algorithm and demonstrate their performance on the test dataset. After optimizing the AUC for each algorithm on the validation dataset, we selected $(\alpha, \beta, \lambda)$ to be $(3, 3, 0)$ for ARCH(c) and SAPBERT, $(3, 1, 0)$ for PubmedBERT and $(1, 1, 0)$ for BioBERT.

### S.6 Additional results

| pairs | type | group | ARCH(c) | ARCH(p) | Pub | Bio | SAP | num |
|---|---|---|---|---|---|---|---|---|
| Code-Code | Similar | PheCode Hierachy | **0.970** | 0.901 | 0.612 | 0.566 | 0.764 | 4094 |
| | | Local Lab Mapping | **0.834** | 0.797 | 0.652 | 0.640 | 0.788 | 1982 |
| | | summary | **0.926** | 0.867 | 0.625 | 0.590 | 0.772 | 6076 |
| | Related | May Treat (Prevent) | **0.797** | 0.791 | 0.630 | 0.586 | 0.587 | 5129 |
| | | Classifies | **0.906** | 0.860 | 0.667 | 0.631 | 0.784 | 4741 |
| | | ddx | **0.776** | 0.747 | 0.610 | 0.568 | 0.634 | 5938 |
| | | Causative | **0.749** | 0.736 | 0.574 | 0.563 | 0.649 | 2873 |
| | | summary | **0.810** | 0.786 | 0.624 | 0.588 | 0.662 | 18681 |
| CUI-Code | Similar | CUI_PheCode | **0.909** | 0.879 | 0.609 | 0.553 | 0.776 | 14096 |
| | | CUI_RXNORM | 0.993 | 0.980 | 0.993 | 0.993 | **0.997** | 1097 |
| | | CUI_LOINC | **0.966** | 0.942 | 0.492 | 0.523 | 0.878 | 165 |
| | | CUI_CCS | **0.982** | 0.958 | 0.875 | 0.790 | 0.972 | 63 |
| | | summary | **0.916** | 0.887 | 0.636 | 0.585 | 0.794 | 15421 |
| CUI-CUI | Similar | Parent | **0.864** | 0.860 | 0.679 | 0.608 | 0.819 | 39374 |
| | | Sibling | 0.857 | **0.879** | 0.688 | 0.570 | 0.743 | 29752 |
| | | summary | 0.861 | **0.869** | 0.683 | 0.592 | 0.786 | 69126 |
| | Related | May Treat (Prevent) | 0.799 | **0.834** | 0.675 | 0.557 | 0.547 | 10593 |
| | | Classifies | **0.918** | 0.897 | 0.660 | 0.583 | 0.826 | 7666 |
| | | ddx | 0.803 | **0.845** | 0.670 | 0.560 | 0.613 | 6062 |
| | | Method_of | **0.900** | 0.871 | 0.509 | 0.512 | 0.734 | 1702 |
| | | Causative | **0.864** | 0.857 | 0.639 | 0.473 | 0.771 | 908 |
| | | summary | 0.843 | **0.857** | 0.658 | 0.559 | 0.661 | 26931 |

Table 5: AUCs of between-vector cosine similarity in detecting known similar pairs and related pairs with 1500-dimensional embedding from ARCH. Within each block, the last column shows the number of known-relation pairs within certain group of pairs.

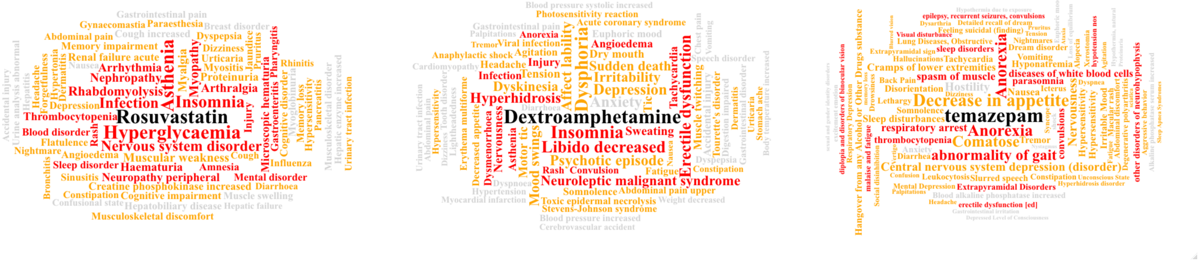


Figure 9: The word cloud of the side effects of three sample drugs - Rosuvastatin, Dextroamphetamine, temazepam. The other words are the description of the side effects. The words colored red are detected by codified only data set while the words colored by orange or red are detected by using both codified data and NLP codes. The words colored by grey are undetected. The size of the words are determined by the cosine similarity with the target drug code.

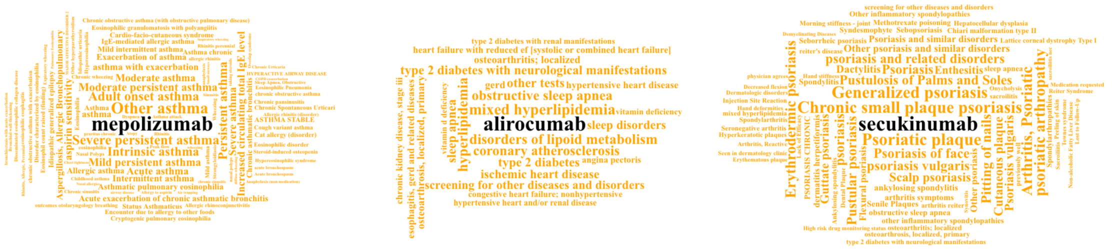


Figure 10: The word cloud of the features detected to be significant related with the three drugs - Mepolizumab, Alirocumab, Secukinumab.
